# Changes in health inequalities following a major urban greenway intervention: Evidence from a 15-year natural experiment in the UK

**DOI:** 10.64898/2026.04.08.26350389

**Authors:** Duyen Nguyen, Ciaran O’Neill, Selin Akaraci, Christopher Tate, Ruoyu Wang, Leandro Garcia, Frank Kee, Ruth F. Hunter

## Abstract

**Highlights:**

- Health inequalities have widened over 15 years, favouring high-income groups
- Inequality in physical activity & mental health widened the most pre-intervention
- Post-intervention, inequalities persisted but stayed relatively unchanged.
- Long-term illness and unemployment were key drivers of inequality
- The greenway may have slowed down the inequality widening but the impact is limited

**Background:** Evidence concerning health inequalities following urban green and blue space UGBS) interventions is limited. This study examined the changes in health inequalities after a major urban regeneration project, the Connswater Community Greenway (CCG), in Belfast, UK.

**Method:** Cross-sectional household surveys were conducted in 2010/11 (baseline), 2017/18 (immediately after completion), and 2023/24 (long-term follow-up) with a sample of approximately 1,000 adults each wave. Using concentration indices (CI), income-related health inequalities for three outcomes (physical activity, mental wellbeing and quality of life) were measured. A regression-based decomposition of concentration index examined the contribution of sociodemographic factors to the observed inequalities underpinning each outcome over time.

**Results:** Across three waves, there was widening of inequalities over the 15-year period across all three health outcomes, with those from high-income groups reported higher levels of physical activity (CI=0.33, SE=0.026), better mental wellbeing (CI=0.03, SE=0.003), and better quality of life (CI=0.09, SE=0.008). The widening inequalities mainly occurred during the construction phase of CCG (2010-2017) and remained stable post-intervention (2017-2023). Decomposition analysis revealed that the pro-poor concentration of long-term illness and unemployment was the key driver that together explained approximately 51%-76% of the inequalities.

**Conclusion:** The CCG was limited in reducing health inequalities which were mainly driven by long-term illness and unemployment - factors beyond the direct scope of the UGBS intervention - resulting in low-income groups likely to fall further behind the wealthier groups. The widening of inequality is consistent with findings from other public interventions that did not have a primary equity focus.

## BACKGROUND

Despite being the focus of public health policy, a substantial body of literature shows that health inequalities have persisted and even widened over time in many high-income countries [1, 2]. In 2020, the UK is experiencing the largest inequalities in life expectancy in a decade, reflected in the unequal pace of health improvement across socio-economic groups [3]. These gaps are widest in large urban areas where problems like poverty, poor housing, and limited access to green and safe outdoor spaces are often compounded [3, 4]. While individual-based interventions can help address the effects of disadvantage, existing evidence emphasises the need of systemic approaches in response to the structural causes of health inequalities stemming from the social determinants of health manifesting in where people are born, grow-up, live, work, and age [1].

The United Nation’s Sustainable Development Goals have highlighted the need to develop healthy and sustainable urban environment that sustain healthy populations and help reduce health inequalities [3, 5]. This initiative highlights the necessity of creating safer neighbourhoods with equitable access to high-quality urban green and blue spaces (UGBS), safe walking and cycling infrastructure, affordable homes, and clean air [3]. To date, there are numerous examples of investments in such interventions globally, but there remains a paucity of evidence on the impact of such interventions on health inequalities [5]. Whilst there is rich literature about the positive association between health and exposure to/use of UGBS, the evidence on the link between access to UGBS and health inequality in the UK is quite scarce [4–6]. A Public Health England review in 2020 focusing on UK setting found that most studies do suggest associations between access to greenspace and health inequality, yet some found no clear relationship [4]. Further evidence synthesis was hindered due to the inconsistent inequality measures and relatively short follow-up time [4].

This study presents an investigation on health inequality over 15-year timeframe in relation to the Connswater Community Greenway (CCG) – a major urban greenway project in Belfast, UK [7]. With £40 million investment, the greenway project was designed to create a 16km continuous cycle and walkway through east Belfast, connecting underused public green spaces, enhanced flood alleviation and rejuvenate the ecosystem along the route of east Belfast’s three primary rivers: Connswater, Loop and Knock rivers [7]. The project was located within the most deprived areas of east Belfast [7, 8] where local communities had an older age profile, high death rate due to cancer, a greater burden of respiratory and other chronic diseases compared to the Northern Ireland average [8].

Focus on potential impact on inequalities, this study aimed to investigate the magnitude of the inequalities across three outcomes: physical activity (PA), mental wellbeing; and quality of life (QoL) before and after the implementation of the greenway. This study also sought to assess how these inequalities changed over time and to identify factors that may have contributed to the persistent health inequalities observed.

## METHOD

### Study setting and data collection

The study site targeted 22 electoral wards (out of 29 wards) in east Belfast whose geographical centroid is within a 1-mile radius of CCG – an area home to about 80,000 residents [9]. Data were collected through repeated cross-sectional surveys in 2010 (the year before the construction of the CCG); 2017 (immediately after the completion and opening of the CCG); and 2023 (six years post-opening). Each wave, approximately 1,000 participants aged ≥16 years were surveyed using random probability sampling strategy. Household addresses were randomly selected using the Royal Mail’s Postal Address File with sampling stratified according to the proportion of the overall population in each target ward. Further details of the survey protocol are reported elsewhere [10].

### Outcomes and covariates

The health inequalities measured in this study focused on three domains of health with known associations with UGBS interventions: PA, mental wellbeing and QoL. PA was measured using the Global Physical Activity Questionnaire [11], with the primary measure being the proportion of the population meeting the recommended PA level per week of at least 150 minutes of moderate-intensity PA or 75 minutes of vigorous-intensity PA or an equivalent combination of the two. Total time spent on PA (converted to an equivalent time spent on moderate-intensity PA in minutes per week) was also examined as a continuous quantitative measure of PA.

Mental wellbeing was examined using the Warwick-Edinburgh Mental Wellbeing Scale (WEMWBS) [12] with the score ranging from 14 to 70 (with a higher score indicative of better overall mental wellbeing). The EQ-5D instrument was used to measure QoL, applying the UK value set of EQ-5D-3L to derive a health-related utility score in waves 1 and 2 [13]. In wave 3 (2023/24) the survey used the E-5D-5L and, as such, a mapping exercise to the EQ-5D-3L [14] was performed to provide a comparable measure for QoL across three waves.

Household income as a five-quintile ranking (from 1 = poorest to 5 = richest) was used as the equity variable of interest for assessing inequalities. The income quintiles were derived from self-reported gross household income, originally collected in broad income brackets that had been adjusted for inflation within each survey wave to ensure comparability over time.

### Statistical analysis

To investigate inequalities in the outcomes of interest, the study used the concentration index to measure and compare the magnitude of inequalities across three waves [15, 16]. Next, a decomposition analysis was conducted to provide further insight into the contribution of several sociodemographic factors toward the inequalities of study outcomes. Lastly, sensitivity analyses were conducted to (i) address missing data issue and (ii) provide an additional comparison using an extended sample covered all 29 wards of east Belfast (the main analysis covered 22 wards located within 1-mile from CCG). All results were weighted to reflect the composition of the whole population by ward-level population size, gender, age and seasonality. All analyses were conducted in STATA 17.0 at a significance level of 5%.

#### Measurement of inequalities

The measurement of inequalities as a concentration index was conceived initially by plotting the cumulative share of an outcome against the population ranked by income status [17]. When the outcome is equally distributed across the whole income spectrum, the concentration curve is positioned at the 45-degree line (the equality line). If a larger share of the outcome came from the poorer group (or the outcome is more prominent among the poor), the concentration curve – connecting the cumulative share of outcome across income distribution from the poorest to the richest – lies above the equality line and vice versa. The concentration index is estimated as twice the area under the equality line to the concentration curve. Together, the concentration curves provided graphical demonstrations of inequality whilst concentration index offered a numerical measurement of inequality across three waves.

Following the literature on inequality measurement, this study used the standard concentration index [17] for continuous outcomes and the Erreygers concentration index for only one binary outcome of PA, measured as the percentage of population meeting the PA recommendation [18]. Income was used as a ranking variable to construct the concentration curves and estimate the concentration indices throughout the study.

#### The decomposition of inequalities

The regression-based decomposition of the concentration index was based on the proposed approach by Wagstaff et al [15] to decompose the inequality in the interested outcome through the inequalities observed in other factors associated to the outcome. The decomposition works by estimating the elasticity (*i.e. how strongly a factor such as education is associated with an outcome such as PA*) and the inequality in the factor of interest across income groups (*i.e. how big the inequality in education by income is*). Taken together across multiple factors, these estimates allow the relative contribution of each factor to inequality in the outcome of interest to be quantified and compared over time.

More formally, the inequality of the outcome can be expressed through multiple factors as: 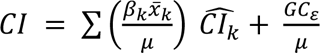 where *CI* is the concentration index of the outcome of interest; 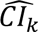 is the concentration index of factor *k*; *x̅*_*k*_ is the average value of factor *k*; *μ* denoting the mean of the outcome and *β*_*k*_ is the average partial effect of factor *k* obtained from the generalised linear model. Here, the contribution of each factor to the inequality in the outcome was the product of its own concentration index 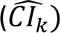 and its elasticity with the outcomes 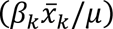, which together constitute the explained part of the inequality in outcome. The *GC*_*ɛ*_ is a generalized concentration index for the error term *ɛ* and 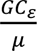 represents the unexplained part of the inequality of the outcome (or the residual).

For the inequalities in percent of people meeting the PA target (as a binary outcome), a modified decomposition was employed: 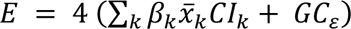 with *E* is Erreygers CI of outcome, *CI*_*k*_ are the standard concentration index of factor *k* and *β*_*k*_obtained from linear probability regression. The contribution of each factor was derived as the percentage share of *β*_*k*_*x̄*_*k*_*CI*_*k*_ in the total inequality (*E*) of the outcome.

Based on previous studies about the association between UGBS and health outcomes [19], a set of factors including gender, age, marital status, education, employment, long-term illness, accommodation types, car ownership and deprivation were used to obtain the correlation between these factors and interested outcomes and how the inequalities in these factors contributed toward the inequalities in the outcomes. Household income was used solely as a ranking variable for constructing the concentration index and not included in the regression for decomposition to avoid double-counting [20]. The regression-based decomposition considered the area-based deprivation variable alongside other factors (housing and car ownership) to capture the effect of economic status on the inequalities of health outcomes.

### Sensitivity analysis

To further validate the result in the main analysis, we extended the investigation using imputed data to overcome potential limitations due to missing data and also a comparative analysis using two additional comparators.

Multiple imputation by chained regression was conducted to impute the missing data on income variable given the substantial missing in this variable (see Supplement 1). Once the imputed data were completed, the measurement and decomposition of inequalities were performed in the same way described above on stacked imputed data [21] to validate the results from the base analysis.

The study also compared changes in health inequalities observed in the present study with two comparative samples: one using Health Survey Northern Ireland (HSNI) [1] and the other employed an extended sample covering all 29 electoral wards of east Belfast (the intervention site covers 22 of these wards). The first comparative sample from HSNI provided deprivation-based inequalities in PA and mental wellbeing of Belfast City and Northern Ireland but only for a single time point (2016/17 for PA and 2017/18 for WEMWBS). The second comparator investigated an extended sample of east Belfast including an additional 200 residents from seven adjacent wards of east Belfast whose geographic centroids lay outside a one-mile radius of the CCG. Although this additional group was relatively small to be a stand-alone comparator [22], its inclusion allowed assessment of whether inequality patterns observed in the intervention area were distinctive or reflected wider trends across east Belfast. Data for the additional group were collected using the same protocols as applied to the main sample, ensuring comparability across the study area [16].

## RESULTS

### 1. Descriptive analysis

Across three waves, the intervention site observed an increase in the percentage of the population aged 36-45 years old and a change in marital status with fewer people married/cohabited (see **Table 1)**. The percentage of the population with higher education reduced slightly while the employment went up at wave 3.

**Table 1:** Participant characteristics across three waves of the household survey.

|  | Percentage |  |  |  | Concentration index (income ranked) |  |  |  |
| --- | --- | --- | --- | --- | --- | --- | --- | --- |
|  | Wave 1<br>N=1037 | Wave 2<br>N=968 | Wave 3<br>N=944 | P-value<sup>a</sup> | Wave 1 | Wave 2 | Wave 3 | P-value<sup>b</sup> |
| **Gender** |  |  |  |  |  |  |  |  |
| Male | 59.0% | 55.5% | 55.4% | 0.18 | 0.07* | 0.04 | 0.08* | 0.09 |
| Female | 41.0% | 44.5% | 44.6% |  |  |  |  |  |
| **Age groups** |  |  |  |  |  |  |  |  |
| 16-25 | 7.6% | 7.1% | 7.1% | 0.87 | -0.25* | -0.17* | -0.07 | 0.62 |
| 26-35 | 18.0% | 18.1% | 18.6% | 0.93 | 0.12* | 0.04 | 0.13* | 0.24 |
| 36-45 | 18.2% | 15.7% | 20.7% | 0.02 | 0.18* | 0.20* | 0.13* | 0.17 |
| 46-55 | 16.5% | 14.6% | 14.3% | 0.34 | 0.11* | 0.15* | 0.12* | 0.62 |
| 56-64 | 13.2% | 16.1% | 14.4% | 0.19 | -0.03 | 0.01 | -0.03 | 0.67 |
| 65+ | 26.4% | 28.4% | 24.9% | 0.22 | -0.33* | -0.34* | -0.31* | 0.22 |
| **Marital status** |  |  |  |  |  |  |  |  |
| Single | 27.8% | 30.9% | 10.7% | <0.001 | -0.16* | -0.14* | 0.29* | <0.001 |
| Married/ Cohabited | 48.8% | 44.7% | 39.2% |  | 0.25* | 0.26* | 0.22* | 0.17 |
| Others | 23.4% | 24.4% | 50.1% |  | -0.36* | -0.36* | -0.24* | <0.001 |
| **Education<sup>c</sup>** |  |  |  |  |  |  |  |  |
| High education | 43.2% | 38.6% | 35.9% | 0.003 | 0.19* | 0.19* | 0.19* | <0.001 |
| Low education | 56.8% | 61.4% | 64.1% |  |  |  |  |  |
| **Employment** |  |  |  |  |  |  |  |  |
| Employed | 52.7% | 50.8% | 58.9% | 0.001 | 0.26* | 0.23* | 0.21* | 0.17 |
| Unemployed/ Others | 47.3% | 49.2% | 41.1% |  |  |  |  |  |
| **Long-term illness** |  |  |  |  |  |  |  |  |
| Yes | 31.0% | 33.0% | 31.0% | 0.56 | -0.27* | -0.31* | -0.34* | <0.001 |
| No | 69.0% | 67.0% | 69.0% |  |  |  |  |  |
| **Accommodation** |  |  |  |  |  |  |  |  |
| Owned | 36.1% | 31.6% | 33.6% | <0.001 | -0.02 | 0.03 | -0.04 | 0.72 |
| Mortgaged | 32.4% | 25.9% | 32.3% |  | 0.36* | 0.40* | 0.35* | 0.29 |
| Rented/ others | 31.5% | 42.5% | 34.1% |  | -0.33* | -0.25* | -0.31* | 0.01 |
| **Car ownership** |  |  |  |  |  |  |  |  |
| No car | 25.5% | 30.3% | 21.0% | <0.001 | -0.44* | -0.43* | -0.47* | 0.049 |
| Only 1 car | 45.7% | 44.1% | 47.7% |  | 0.02 | 0.03 | -0.03 | 0.03 |
| 2+ cars | 28.8% | 25.6% | 31.4% |  | 0.40* | 0.43* | 0.35* | 0.23 |
| **Deprivation** |  |  |  |  |  |  |  |  |
| 1 (most deprived) | 22.8% | 23.3% | 21.7% | <0.001 | -0.27* | -0.29* | -0.29* | 0.02 |
| 2 | 17.8% | 18.7% | 13.6% |  | -0.07 | -0.09 | -0.12 | 0.67 |
| 3 | 22.7% | 19.7% | 12.0% |  | 0.06 | -0.02 | 0.03 | 0.95 |
| 4 | 18.4% | 21.9% | 20.1% |  | 0.22* | 0.21* | -0.02 | <0.001 |
| 5 (least deprived) | 18.3% | 16.3% | 32.6% |  | 0.24* | 0.29* | 0.25* | 0.46 |

\* Wave 1: 2010, baseline; Wave 2: 2017/2018 – one year post the opening of CCG; Wave 3: 2023/2024 long term follow-up at 60year post opening. a – p value obtained from Pearson's chi-square tests, b - indicate standard concentration index is significantly different from 0 (significant inequality) at p<0.05, c – high education (at least obtained apprentice/ A-level or a degree) vs low (GCSE or lower)

In regard to inequality, consistently throughout the three waves, the high-income group comprised a larger proportion of individuals in the productive age range (26 –55 years old) and demonstrated a higher rate of employment and higher educational attainment. In contrast, older adults (aged ≥65 years) and those with long-term illnesses were significantly more concentrated among low-income groups. Less-affluent individuals were also more likely to live in rented accommodation and not have a car, reflecting multiple dimensions of socioeconomic disadvantage among the low-income group.

### 2. Measurement of health inequality

The results indicated that there were persistent and widening inequalities presented in PA, mental wellbeing and QoL, with better outcomes concentrated more among high-income groups (see **Table 2**). However, the increase inequality was most pronounced pre-intervention, followed by a modest, non-significant increase during post-intervention period (2017 vs 2023).

**Table 2:** Income-related inequalities in physical activity, mental wellbeing and quality of life.

| Indicators | Concentration index (SE) |  |  | Significant change (p-value) <sup>+</sup> |  |  |
| --- | --- | --- | --- | --- | --- | --- |
|  | Wave 1<br>(2010) | Wave 2<br>(2017) | Wave 3<br>(2023) | Wave<br>1 vs 2 | Wave<br>2 vs 3 | Wave<br>1 vs 3 |
| Meet the PA target <sup>a</sup> | 0.137*<br>(0.036) | 0.303*<br>(0.041) | 0.335*<br>(0.026) | ↗<br>P=0.002 | ↔<br>P=0.522 | ↗<br>P<0.001 |
| Time spent in PA <sup>b</sup> | -0.001<br>(0.030) | 0.122*<br>(0.029) | 0.130*<br>(0.024) | ↗<br>P=0.004 | ↔<br>P=0.831 | ↗<br>P=0.001 |
| Mental wellbeing<br>(WEMWBS) <sup>b</sup> | 0.022*<br>(0.003) | 0.032*<br>(0.004) | 0.034*<br>(0.003) | ↔<br>P=0.061 | ↔<br>P=0.666 | ↗<br>P=0.017 |
| Quality of life<br>(EQ5D3L) <sup>b</sup> | 0.066*<br>(0.006) | 0.066*<br>(0.008) | 0.088*<br>(0.008) | ↔<br>P=0.995 | ↔<br>P=0.054 | ↗<br>P=0.032 |

\* significant inequality at $p < 0.05$ . A positive CI means the outcome is more concentrated among the rich and vice versa. <sup>+</sup> p-value was provided to show the significant change in inequalities, with ↗ indicating significant increase and ↔ means no significant change at $p < 0.05$ . a – Erreygers concentration index was reported; b - standard concentration index was reported.

First, in term of population share meeting the PA recommendation, the estimated inequality doubled during the pre-intervention period from CI_2010_ = 0.137 (SE = 0.036) in 2010 to CI_2017_ = 0.303 (SE = 0.041; p_2010 vs 2017_ = 0.002) in 2017 and remained at the same level during the post-intervention period (CI_2023_ = 0.335, SE = 0.026) (p_2017 vs 2023_ = 0.522). Similar pattern of inequality change was also seen in the concentration indices of the quantitative measure as total time of PA per week.

The concentration curves for two measures of PA showed a progressive drift from the equality line over time but the two curves in 2017 and 2023 was almost overlapped indicating a similar level of inequalities in PA post-intervention (see **Figure 1**). This result was reiterated in the sensitivity analysis with imputed data (see Supplement 1) and with the extended sample of east Belfast (see Supplement 2). Collectively, the results demonstrate a persistent and widening inequality in PA over time in favour of the high-income groups, either in terms of meeting the recommended PA guidelines or time spent in PA.

**Figure 1:**
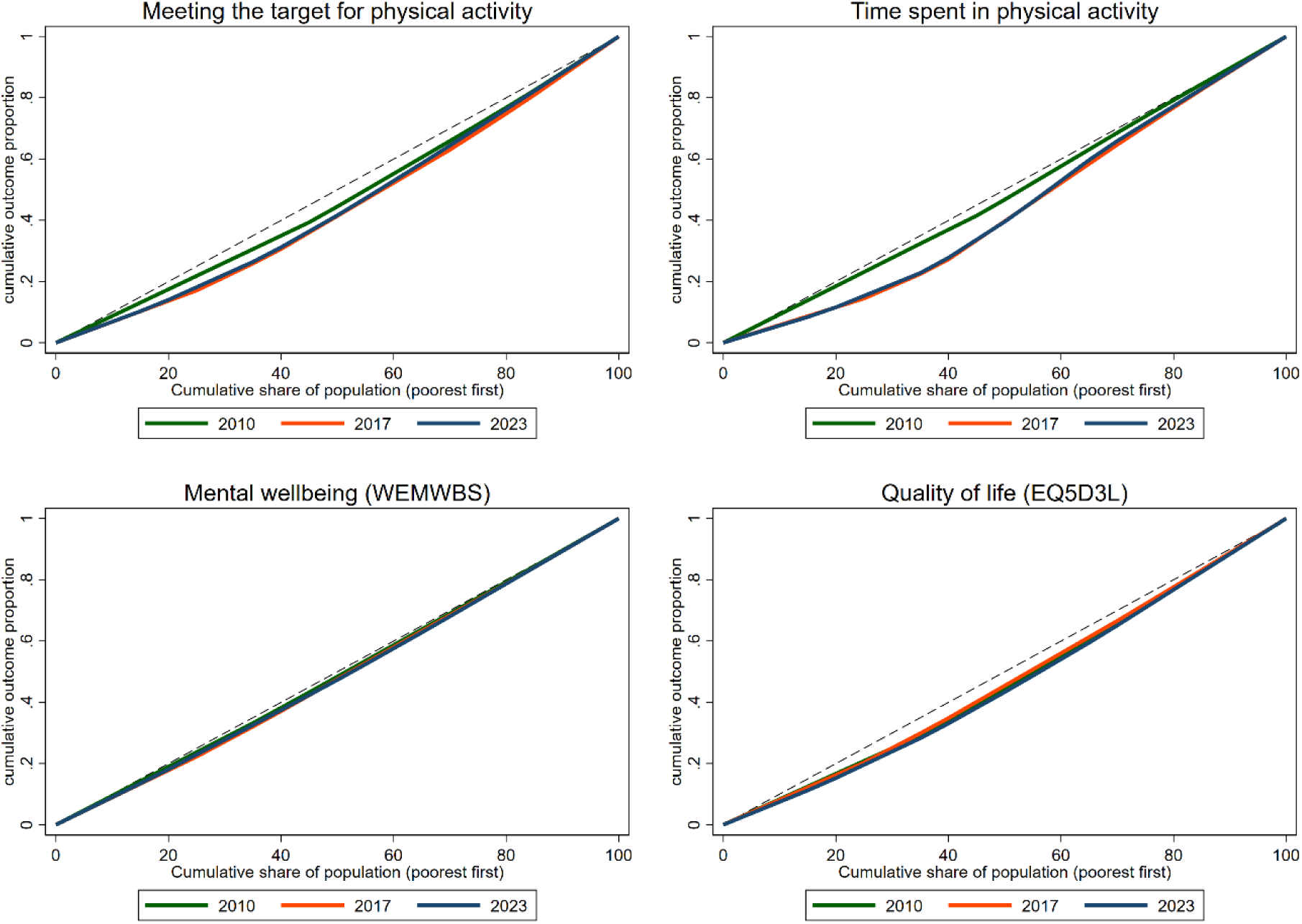
Concentration curves of physical activity, mental wellbeing and quality of life.

The inequality in mental wellbeing was the smallest among the three outcomes but demonstrated a similar distribution of higher mental well-being concentrated among high-income individuals. The CI at baseline was CI_2010_ = 0.022 (SE = 0.003) and increase to CI_2017_ = 0.032 (SE = 0.004) by 2017 with a marginal statistical significance (p_2010 vs 2017_ =0.061), whilst the inequality during post-intervention period (2017-2023) remained largely unchanged (CI₂₀₂₃ = 0.034; p_2017 vs 2023_ = 0.666). Comparative results from HSNI and the extended sample of east Belfast (see Supplement 2) and the imputed data also shared a similar direction.

QoL showed similar inequalities that favoured high-income individuals, but the pattern of change was slightly different from PA and mental wellbeing. During pre-intervention period, there was no change in the inequality measure (CI=0.066 in both 2010 and 2017, p_2010 vs 2017_ = 0.995). The post-intervention period, in contrast, showed an increase in inequality from CI_2017_ = 0.066 to CI_2023_ = 0.088 with the difference being borderline significant with *p_2017 vs 2023_* = 0.054. The sensitivity analysis supports the observation of an increasing trend in inequality in QoL between 2017 and 2023 since the inequality in QoL in the two time points were shown statistically increase in both the imputed dataset (*p* = 0.007) and the extended sample (*p* = 0.047).

### 3. Decomposition of inequalities

**Figure 2** shows the percentage contribution of several sociodemographic factors to the inequalities in each outcome, and how the pattern of contributions changed across the three waves. Long-term illness and employment both consistently contributed to inequalities across each outcome in the three waves. Detailed information of the inequalities of each factor and its elasticity with the outcomes is reported in Supplement 3.

**Figure 2:**
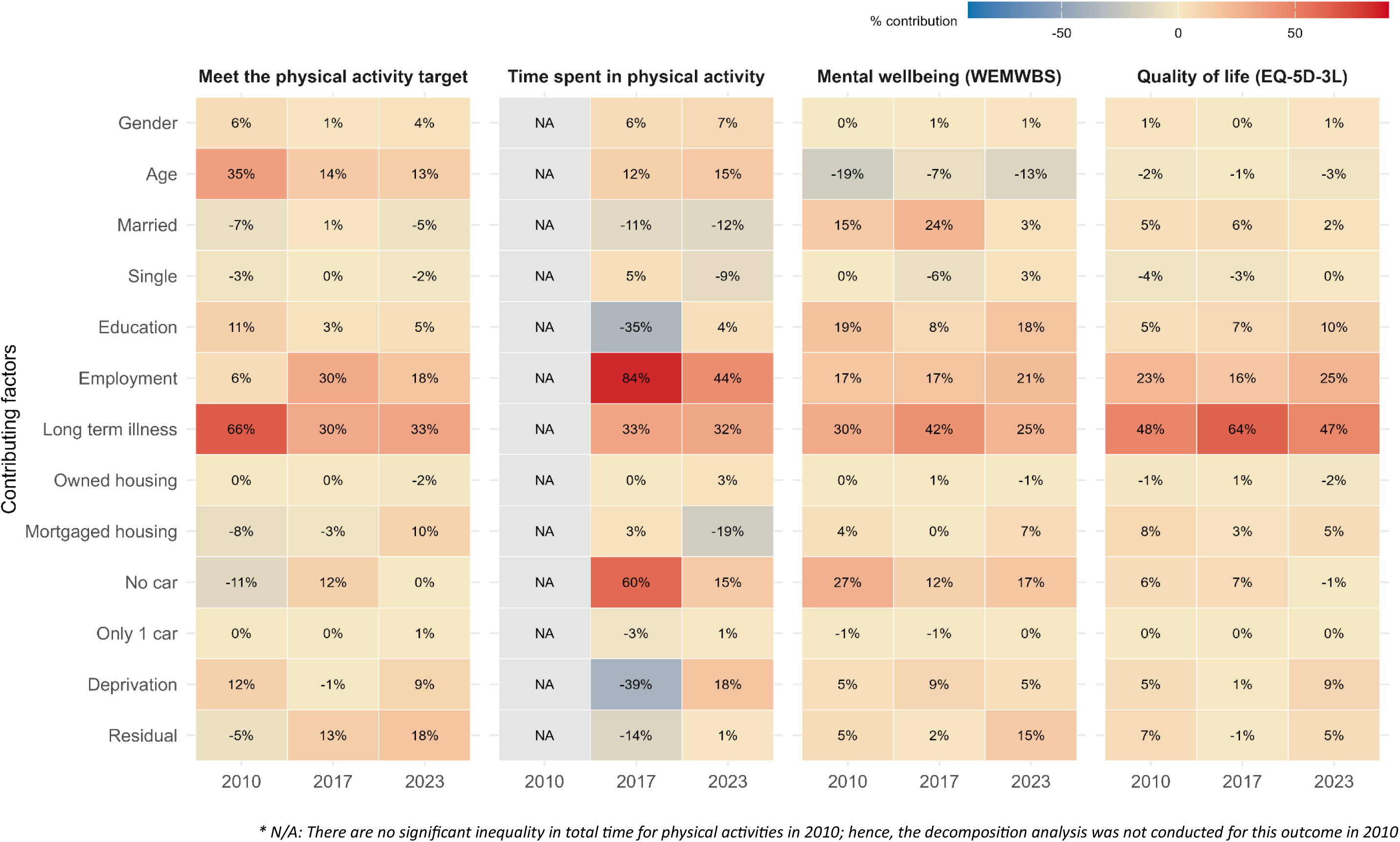
The percentage contribution of covariates in the inequalities of study outcomes.

For PA, long-term illness accounted for up to 66% of total inequality at baseline and remained a major contributor in later waves – 30% in 2017 and 33% in 2023 – indicating the persistence of health-related inequalities acting as a barrier to PA. As expected, long-term illness was more prevalent among lower-income groups (CI= −0.267, see Supplement 3). Employment emerged as the second-largest contributor to the inequality in PA in later waves (30% in 2017 and 18% in 2023) and this can largely be attributed to the positive correlation between employment and PA.

Comparable contributions to inequalities in time spent in PA were also attributable to employment and long-term illness. Interestingly, we noted a 60% contribution to time spent in PA inequality from not owning a car in 2017, indicating that opportunities for PA were potentially restricted to those with the means to travel (the largest negative elasticity was - 0.169 across three waves). This can be explained by low-income groups being significantly less likely to own a car. However, by 2023, the presence of the CCG seemed to weaken the earlier link of not owning a car and lower PA.

Long-term illness also contributed significantly to inequalities in mental wellbeing, explaining approximately 30%, 42% and 25% of the observed disparity in mental wellbeing across the three waves, respectively. Employment was another contributor with steady contributions of around 17%, 17%, and 21% of the total inequality of mental wellbeing across the three waves. The decomposition also identified a negative impact of long-term illness on QoL, which accounted for 47% to 64% of the total variation in QoL. Employment ranked as the second-largest contributor, although its contribution was smaller, ranging from 16% to 25%.

The sensitivity analysis using imputed data also demonstrated a similar pattern (see Supplement 1), reinforcing the roles of long-term illness and employment in the inequality observed in PA, mental health and QoL.

## DISCUSSION

### The pro-rich concentration of health inequalities

Across each of the three studied outcomes, the income gradient was persistently observed to be in favour of the least deprived, with a higher population share meeting the PA recommendation, more time spent in PA, better mental wellbeing and higher QoL. This finding echoes the results of earlier studies conducted since the early 2000s and 2010s [23–25]. For example, a study conducted in England using data from the Health Survey for England in 2008-2016 and the Active People Survey (2004-11) [24] reported a consistent pro-rich inequality in PA, indicating that higher socioeconomic status groups were overall more physically active and accumulated more time on moderate-to-vigorous PA. However, a systematic review of European studies found no substantial inequalities in time spent on PA during 2000-2010, similar to the non-significant inequalities in time for PA observed in 2010 in our study [25].

Socioeconomic disparities were also observed for mental wellbeing in line with other studies across Europe and UK with different measurements and databases [26–28]. Thomson et al investigated UK national survey data from 1991 to 2014 and found that socioeconomic inequalities in general health were consistently more concentrated among the poor and increased after 2010 [26]. A study in nine high-income European countries from 2002-2015 also observed a high concentration of depressive symptoms among lower SES [27].

The evidence of pro-rich inequality in QoL was aligned with large-scale analyses in the UK and neighbouring countries which also reported worsen QoL on average in more deprived groups as compared to more affluent communities [29, 30]. Population studies across Europe also pointed to a highly persistent phenomenon of inequality in mortality and mortality with relative inequalities have almost universally increased over the past decade despite reducing inequalities has been the focus of public health policy in many countries [31].

### The direction of change in health inequalities

Over 15 years of observation, we found that inequalities with respect to PA, mental wellbeing and QoL have widened significantly, however, the direction of inequalities change was evidently different pre- and post-intervention.

For PA and mental wellbeing, an increase in inequalities was observed during pre-intervention (2010 vs 2017) but stayed relatively stable post-intervention (2017 vs 2023). The increase in inequalities was the most evident for PA, which is similar to the broader trend of widening inequalities across the UK and Europe during the same period [2, 3, 24, 25]. During the post-intervention period, we observed a plateauing of inequalities in PA and mental wellbeing, which coincides with the presence of the CCG. This temporal overlap suggests a potential influence of greenways that might have mitigated further divergence of changes (particularly in PA and mental wellbeing) across different socioeconomic classes. Due to a lack of comparable evidence on whether the inequalities in PA and mental wellbeing are widening or narrowing during 2017-2024 in the UK, further research is warranted to test if the stabilisation of inequalities observed here is attributable to the CCG or reflects broader structural changes.

The present study also found a different trajectory of inequality change with respect to QoL, which showed widening inequalities in the post-intervention period after remaining relatively stable pre-intervention. This was likely due to the multi-dimensional characteristic of the EQ-5D measurement capturing multiple domains of health, hence it might be less sensitive to environmental changes. When both low-income and high-income groups experience comparable gains from a community-level intervention such as the CCG, then overall QoL may improve, but the relative inequality may remain largely unchanged.

Regardless of the difference in inequality change pre- and post-intervention, the overall trajectory of health inequalities from 2010 (baseline) to 2023 (five years post-intervention) was upward. This result aligns with the wider trend of worsening health inequalities in the UK [2, 3], and yet represents another example of the theory of diffusion [32]. This phenomenon was often seen after population-based interventions or policies, as healthier behaviours are typically adopted first by higher socioeconomic status groups leading a (temporarily) widening social gaps before benefits eventually diffuse through the population [3, 32].

### Decomposition of inequalities in physical activity, mental wellbeing and quality of life

The decomposition analysis revealed that long-term illness and employment were the leading contributors to inequalities observed in PA, mental wellbeing and QoL. Although there are limited comparable studies in terms of methodology, this result was consistent with previous inequality-focused studies in the UK and other high-income countries [24, 27, 33]. For example, McElroy et al [27] found that high rate of unemployment and ill-health in low-income groups drove much of the socioeconomic gradient in mental wellbeing across nine European countries. Gunasekara et al [33] also reported that unemployment was one of the three biggest contributors to health inequalities in Australia and New Zealand. For PA, we did not find similar studies in the UK; however, there are a few studies that have identified income and education as primary predictors of PA, while employment and health status often provide indirect effects [24].

This finding has added to the growing evidence pool of health inequalities pointing toward the role of underlying social stratification – here particularly referred to the socioeconomic gradient in long-term illness and unemployment – in addressing health inequality. As the inequality in these determinants persisted over time, it is to be expected that inequalities in physical activity, health, and wellbeing would also remain. Yet the plateauing of inequality in PA and mental wellbeing post-intervention may indicate that neighbourhood-level improvements in the most deprived areas may buffer, but not eliminate, the inequalities in healthy behaviours and overall health.

### Strengths and limitations

This study is one among very few investigating the trajectory of health inequalities across multiple health outcomes following an UGBS intervention. The use of empirical data collated over 15 years before and after the intervention along with the decomposition analysis has provided useful insights into the drivers of inequality over time. It also adds new evidence to the limited post-COVID-19 literature since the majority of current literature was based on data from the early 2000s to mid-2010s, and lacks population-based evidence post-2020 [2, 4, 23].

However, there are certain limitations that should be noted for further application of our study results. First, the study used data from a repeated cross-sectional data set with three time points spanning almost 15 years; hence, the time trends captured in our study are relative comparisons across survey waves rather than reflecting a continuous temporal pattern. This also limited the exclusion of the COVID-19 impact on overall health and health inequalities Secondly, data were collected in an urban setting where only 5% of Northern Ireland’s total population resided, hence, the findings reflect the local social and demographic context of east Belfast and may not be generalisable to other urban areas. Additionally, the income variable was not equivalised for household size, it holds certain limitation to represent the true economic status for participants and could pose bias in the measure of inequalities.

Thirdly, although the decomposition technique is useful in breaking down the health inequalities into a more versatile combination of structural inequalities in social determinants, drawing causal inferences about what drives changes in health inequalities from decomposition analysis is challenging. Regression-based decomposition is conceptually based on associations between the outcome and a set of explanatory factors rather than on causal identification. As such, the findings indicate which factors are associated with the observed patterning of inequality, but do not establish whether those factors independently caused it. This is further complicated by the fact that the current study exploited a natural experiment that does not correspond to well-defined intervention assumptions, hence, the findings should not be interpreted in overly strong causal terms.

## CONCLUSION

This study is the first study in the UK and one of very few studies internationally to examine socioeconomic inequalities in multiple health outcomes before and after the implementation of a UGBS intervention spanning over 15 years of observation. Inequalities in PA and mental wellbeing were consistently favourable of high-income groups with a noticeable increasing tendency between 2010-2017 (pre-intervention) but stabilised at long-term follow-up period coinciding with the presence of greenway in the community. The decomposition analysis revealed that the poor-concentrated distribution of long-term illness and unemployment can explain a majority of the persistent inequalities observed in PA, mental wellness and QoL. Together, these findings suggest that the CCG may have helped to stabilise the widening inequalities in PA and mental wellbeing in post-intervention, but the impacts were rather limited since the broader causes of inequalities were stemmed from long-term illness and unemployment leaving low-income groups unable to keep pace with the rich in achieving more PA and better mental wellbeing.

## Data Availability

The datasets used and/or analysed during the current study are available from the corresponding author on reasonable request.

## Acknowledgement

GroundsWell is an interdisciplinary consortium involving researchers, policy, implementers and communities. It is led by Queen’s University Belfast, University of Edinburgh and University of Liverpool in partnership with Cranfield University, University of Exeter, University of Glasgow, University of Lancaster and Liverpool John Moores University.

We would like to acknowledge our stakeholders including: Belfast, Edinburgh and Liverpool City Councils, Public Health Agencies of Scotland and Northern Ireland, Greenspace Scotland, Scottish Forestry, Edinburgh and Lothians Health Foundation, Department for Infrastructure Northern Ireland, Belfast Healthy Cities, Climate Northern Ireland, Health Data Research UK, Administrative Data Research Centre, NatureScot, Mersey Care NHS Foundation Trust, Liverpool City Region Combined Authority, Liverpool Health Partners, NHS Liverpool Clinical Commissioning Group, the Scottish Government, Edinburgh Health and Social Care Partnership, HSC Research and Development Office Northern Ireland, EastSide Partnership, Ashton Centre, Regenerus, Sustrans, Cycling UK, CHANGES, The Mersey Forest, Translink, Anaeko, AECOM, The Paul Hogarth Company and Moai Digital.

We also wish to acknowledge the co-investigators involved in the PARC Study (which collected the data on the wave 1 and wave 2 household surveys, and other elements of the protocol development): Mark A Tully, Helen McAneney, Margaret E Cupples, Michael Donnelly, George Hutchinson, Alberto Longo, Lindsay Prior, Michael Stevenson. The PARC study (which funded wave 1 and wave 2 household survey data collection) is supported by a grant from the National Prevention Research Initiative. The Funding Partners are (in alphabetical order): Alzheimer’s Research Trust; Alzheimer’s Society; Biotechnology and Biological Sciences Research Council; British Heart Foundation; Cancer Research UK; Chief Scientist Office, Scottish Government Health Directorate; Department of Health; Diabetes UK; Economic and Social Research Council; Engineering and Physical Sciences Research Council; Health and Social Care Research and Development Division of the Public Health Agency (HSC R&D Division); Medical Research Council; The Stroke Association; Welsh Assembly Government and World Cancer Research Fund.

In addition, the authors would like to acknowledge the partners and stakeholders involved in the PARC Study, including: Wendy Langham, CCG Manager; East Belfast Partnership; Belfast City Council; Department of Health, Social Services and Public Safety; Department for Regional Development; Department of the Environment; Department for Social Development; Belfast Health and Social Care Trust; East Belfast Community Development Agency; SportNI; Belfast Healthy Cities; Sustrans; Public Health Agency; Ordnance Survey NI and the local residents of the CCG population. Part of this work were presented as an oral presentation at the Prevention Research 2026 in Birmingham (UK) in March 2026.

## Declaration of interests

### Ethical approval

Ethical approval for Wave 1 and Wave 2 was obtained from the Office for Research Ethics Committees Northern Ireland (09/NIR02/66). Additional approval for the Wave 3 survey was granted by the Medicine, Health and Life Sciences Research Ethics Committee (MHLS 22_161 – Amendment 1).

### Consent for publication

Participants provided written informed consent prior to taking part in the study.

### Patient and public involvement (PPI statement)

Patients and/or the public were involved in the design, or conduct, or reporting, or dissemination plans of this research. Refer to the protocol paper for further details (available at https://doi.org/10.1136/bmjopen-2024-097530).

### CRediT author statement

**Duyen Nguyen:** Conceptualization, Methodology, Formal analysis, Visualization, Writing - Original Draft

**Ciaran O’Neill:** Methodology, Writing - Review & Editing, Funding acquisition

**Selin Akaraci:** Writing - Review & Editing

**Christopher Tate:** Writing - Review & Editing

**Ruoyu Wang:** Writing - Review & Editing

**Leandro Garcia:** Writing - Review & Editing

**Frank Kee:** Writing - Review & Editing, Project administration, Funding acquisition

**Ruth Hunter:** Conceptualization, Investigation, Methodology, Writing - Review & Editing, Supervision, Project administration, Funding acquisition

## REFERENCES

1. Rad J: Health inequities: a persistent global challenge from past to future. Int J Equity Health 2025, 24(1):148.

2. Holdroyd I, Vodden A, Srinivasan A, Kuhn I, Bambra C, Ford JA: Systematic review of the effectiveness of the health inequalities strategy in England between 1999 and 2010. BMJ Open 2022, 12(9):e063137.

3. Marmot M: Health equity in England: the Marmot review 10 years on. BMJ 2020, 368:m693.

4. Public Health England: Improving Access to Greenspace: A New Review for 2020. In. London, UK: Public Health England; 2020.

5. Hunter RF, Cleland C, Cleary A, Droomers M, Wheeler BW, Sinnett D, Nieuwenhuijsen MJ, Braubach M: Environmental, health, wellbeing, social and equity effects of urban green space interventions: A meta-narrative evidence synthesis. Environment International 2019, 130:104923.

6. WHO Regional Office for Europe: Urban green spaces and health — A review of evidence. In. Copenhagen: World Health Organization Regional Office for Europe; 2016.

7. EastSide Partnership: The Connswater Community Greenway. 2025.

8. Northern Ireland Neighbourhood Information Service (NINIS): Constituency Profile: Belfast East (September 2010). In. Belfast: Northern Ireland Assembly; 2010.

9. Northern Ireland Statistics Research Agency: Northern Ireland Census 2021. In.: NISRA; 2022.

10. Hunter RF, Cleland C, Wang R, O’Neill C, Mullineaux S, Tate C, Küçükali H, Akaraci S, O’Kane N, Garcia L et al: Investigating the long-term public health and co-benefit impacts of an urban greenway intervention in the UK: a natural experiment evaluation - study protocol. BMJ Open 2025, 15(7):e097530.

11. World Health Organization: Global Physical Activity Questionnaire (GPAQ): Analysis guide. In.: WHO; 2002.

12. Tennant R, Hiller L, Fishwick R, Platt S, Joseph S, Weich S, Stewart-Brown S: The Warwick-Edinburgh Mental Well-being Scale (WEMWBS): Development and UK validation. Health and Quality of Life Outcomes 2007, 5:63.

13. Mulhern B, Feng Y, Shah K, Janssen MF, Herdman M, van Hout B, Devlin N: Comparing the UK EQ-5D-3L and English EQ-5D-5L Value Sets. PharmacoEconomics 2018, 36(6):699–713.

14. Hernández-Alava M, Pudney S: Eq5Dmap: A Command for Mapping between EQ-5D-3L and EQ-5D-5L. The Stata Journal 2018, 18(2):395–415.

15. O’Donnell O, van Doorslaer E, Wagstaff A, Lindelow M: Analyzing Health Equity Using Household Survey Data: A Guide to Techniques and Their Implementation. Washington, DC: World Bank; 2008.

16. O’Donnell O, O’Neill S, Van Ourti T, Walsh B: conindex: Estimation of concentration indices. Stata J 2016, 16(1):112–138.

17. Kakwani N, Wagstaff A, Van Doorslaer E: Socioeconomic inequalities in health: measurement, computation, and statistical inference. Journal of econometrics 1997, 77(1):87–103.

18. Erreygers G: Correcting the concentration index. Journal of health economics 2009, 28(2):504–515.

19. Hunter RF, Adlakha D, Cardwell C, Cupples ME, Donnelly M, Ellis G, Gough A, Hutchinson G, Kearney T, Longo A et al: Investigating the physical activity, health, wellbeing, social and environmental effects of a new urban greenway: a natural experiment (the PARC study). Int J Behav Nutr Phys Act 2021, 18(1):142.

20. Erreygers G, Kessels R: Regression-Based Decompositions of Rank-Dependent Indicators of Socioeconomic Inequality of Health. In: Health and Inequality. vol. 21: Emerald Group Publishing Limited; 2013: 0.

21. Nguyen DT, Donnelly M, Van Hoang M, O’Neill C: The case for individualised public health interventions: Smoking prevalence and inequalities in Northern Ireland 1985-2015. Health Policy 2023, 135:104879.

22. Ataguba JE: A short note revisiting the concentration index: Does the normalization of the concentration index matter? Health Economics 2022, 31(7):1506–1512.

23. Scholes S, Mindell JS: Inequalities in participation and time spent in moderate-to-vigorous physical activity: a pooled analysis of the cross-sectional health surveys for England 2008, 2012, and 2016. BMC Public Health 2020, 20(1):361.

24. Farrell L, Hollingsworth B, Propper C, Shields MA: The socioeconomic gradient in physical inactivity: Evidence from one million adults in England. Social Science & Medicine 2014, 123:55–63.

25. Beenackers MA, Kamphuis CBM, Giskes K, Brug J, Kunst AE, Burdorf A, van Lenthe FJ: Socioeconomic inequalities in occupational, leisure-time, and transport related physical activity among European adults: A systematic review. International Journal of Behavioral Nutrition and Physical Activity 2012, 9(1):116.

26. Thomson RM, Niedzwiedz CL, Katikireddi SV: Trends in gender and socioeconomic inequalities in mental health following the Great Recession and subsequent austerity policies: a repeat cross-sectional analysis of the Health Surveys for England. BMJ Open 2018, 8(8):e022924.

27. McElroy B, Walsh E: A happy home? Socio-economic inequalities in depressive symptoms and the role of housing quality in nine European countries. BMC Public Health 2023, 23(1):2203.

28. Tibber MS, Walji F, Kirkbride JB, Huddy V: The association between income inequality and adult mental health at the subnational level-a systematic review. Social psychiatry and psychiatric epidemiology 2022, 57(1):1–24.

29. Szende A, Janssen MF, Cabases J, Ramos-Goni JM, Burström K: Socio-demographic indicators of self-reported health based on EQ-5D-3L: A cross-country analysis of population surveys from 18 countries. Frontiers in public health 2022, 10:959252.

30. Schneider P, Love-Koh J, McNamara S, Doran T, Gutacker N: Socioeconomic inequalities in HRQoL in England: an age-sex stratified analysis. Health Qual Life Outcomes 2022, 20(1):121.

31. Vineis P, Avendano-Pabon M, Barros H, Bartley M, Carmeli C, Carra L, Chadeau-Hyam M, Costa G, Delpierre C, D’Errico A et al: Special Report: The Biology of Inequalities in Health: The Lifepath Consortium. 2020, Volume 8 - 2020.

32. Frohlich KL, Potvin L: Transcending the known in public health practice: the inequality paradox: the population approach and vulnerable populations. American journal of public health 2008, 98(2):216–221.

33. Gunasekara FI, Carter K, McKenzie S: Income-related health inequalities in working age men and women in Australia and New Zealand. Australian and New Zealand Journal of Public Health 2013, 37(3):211–217.

