## Supplementary file for "Changes in health inequalities following a major urban greenway intervention: Evidence from a 15-year natural experiment in the UK"

### Supplement 1: Sensitivity analysis with imputed data

The missing income variable varied across three waves but was relatively substantial in the combined set (wave 1: 13.4%, wave 2: 20.4%, and wave 3: 4.9%). Table S6 below showed the likelihood of missingness in income base on individuals' characteristics. As can be seen, there is significant correlation between marital status, employment, deprivation and survey wave and the missingness of income variable (p<0.05), hence, the pattern of missing in income is likely to be predicted based on these variables (missing-at-random). Therefore, a multiple imputation by chained equations (m=20 imputations) was conducted using the selected covariates in table below to impute the missing data on income. After imputed, the magnitude and changes of inequalities was represented in Table S7, with the decomposition analysis shown in Figure S1.

**Table S1: Pattern of missingness**

| Covariates | Coefficient | 95% confidence interval | p-value |
|---|---|---|---|
| Gender | -0.002 | -0.027 to 0.023 | 0.876 |
| Age | -0.001 | -0.002 to 0.000 | 0.056 |
| Survey wave | -0.005 | -0.007 to -0.003 | <0.001 |
| Marital status | -0.024 | -0.042 to -0.005 | 0.011 |
| Education | -0.010 | -0.038 to 0.017 | 0.454 |
| Employment status | -0.087 | -0.117 to -0.058 | <0.001 |
| Area-based deprivation | 0.012 | 0.003 to 0.021 | 0.007 |

**Table S2: Concentration index of interested outcomes (estimated with imputed data)**

| | Concentration index (SE) | | | Significant change ** | | |
|---|---|---|---|---|---|---|
| Indicators | 2010 | 2017 | 2024 | Wave 1 vs 2 | Wave 2 vs 3 | Wave 1 vs 3 |
| Meet PA target | 0.139* (0.034) | 0.301* (0.035) | 0.326* (0.026) | ↗ | ↔ | ↗ |
| Time spent in PA | 0.006 (0.027) | 0.125* (0.026) | 0.121* (0.023) | ↗ | ↔ | ↗ |
| Mental wellbeing (WEMWBS) | 0.021* (0.003) | 0.031* (0.003) | 0.033* (0.003) | ↗ | ↔ | ↗ |
| Quality of life (EQ5D3L) | 0.061* (0.005) | 0.060* (0.006) | 0.087* (0.008) | ↔ | ↗ | ↗ |

* Significant inequality with p<0.05. A positive CI means the outcome is more concentrated among the rich, a negative CI means the outcome is more concentrated among the poor. ** Significant difference in the estimated inequalities of two waves detected at p<0.05 using z-test

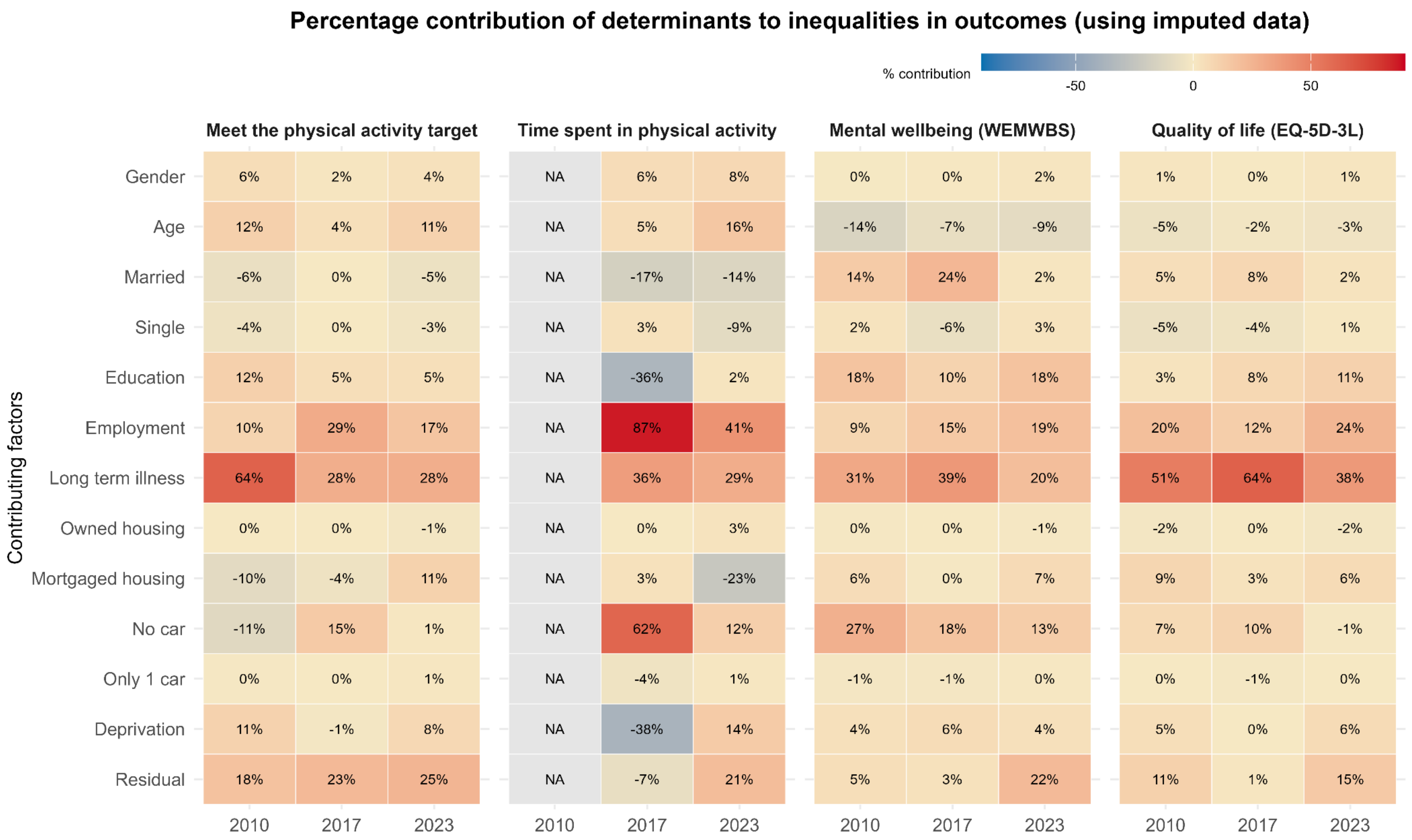

**Figure S1: The percentage contribution of covariates in the inequalities of study outcomes (using imputed data)**

### Supplement 2: Health inequalities in the extended sample of east Belfast, wider Belfast and Northern Ireland

**Table S3: Inequalities of different indices of East Belfast [a] (extended sample) across three waves**

| EAST BELFAST [a] | Concentration index (CI) | | | Significant change [b] | | |
|---|---|---|---|---|---|---|
| | 2010 | 2017 | 2024 | Wave 1 vs 2 | Wave 2 vs 3 | Wave 1 vs 3 |
| Meeting PA target | 0.142* (0.032) | 0.266* (0.036) | 0.284* (0.025) | ↗ | ↔ | ↗ |
| Time spent in PA | 0.008 (0.024) | 0.104* (0.025) | 0.109* (0.022) | ↗ | ↔ | ↗ |
| WEMWBS score | 0.019* (0.003) | 0.032* (0.003) | 0.033* (0.003) | ↗ | ↔ | ↗ |
| Quality of life (EQ5D3L) | 0.063* (0.005) | 0.056* (0.007) | 0.082* (0.007) | ↔ | ↗ | ↗ |

* Significant inequality with p<0.05. a- The results were estimated for all 29 wards in East Belfast while the main analysis focused on 22 wards whose geographical centroid of within 1-mile radius of CCG. b- A significant difference in the inequalities of two waves at p<0.05 using z-test. A positive CI means the outcome is more concentrated among the rich, a negative CI means the outcome is more concentrated among the poor

We also obtained population data across Belfast (county-level) and Northern Ireland from the Health Survey Northern Ireland (HSNI) [1] for additional comparison of the estimated inequalities in PA and mental wellbeing in the present study (quality of life data not available in these comparator samples). The HSNI data provided estimates of deprivation-based inequalities in PA and WEMWBS of Belfast and Northern Ireland, although data were available only for a single time point (2016/17 for PA and 2017/18 for WEMWBS). For PA, although HSNI and the current study used two slightly different tool, literature showed that the two questionnaires provided moderate to high validity [2].

**Table S4: Deprivation-related inequalities in PA and mental wellbeing using HSNI data [1]**

| Year | Source | Area | N* | Concentration index ** | SD | p-value |
|---|---|---|---|---|---|---|
| **Meeting the PA target (current study used GPAQ; HSNI used IPAQ)*** ** | | | | | | |
| 2016/17 | HSNI | Belfast | 349 | 0.079 | 0.031 | 0.011 |
| 2016/17 | HSNI | Northern Ireland | 1908 | 0.049 | 0.012 | 0.0001 |
| 2017/18 | Current study | CCG (intervention site) | 968 | 0.011 | 0.013 | 0.399 |
| 2017/18 | Current study | East Belfast | 1214 | 0.021 | 0.011 | 0.069 |
| **Mental wellbeing (WEMWBS)** | | | | | | |
| 2017/18 | HSNI | Belfast | 479 | 0.018 | 0.005 | <0.001 |
| 2017/18 | HSNI | Northern Ireland | 2922 | 0.010 | 0.002 | <0.001 |
| 2017/18 | Current study | CCG (intervention site) | 968 | 0.017 | 0.003 | <0.001 |
| 2017/18 | Current study | East Belfast | 1214 | 0.016 | 0.003 | <0.001 |

* N= the sample size from the corresponding survey. ** Erreygers CI was provided for physical activity, standard CI for WEMWBS. ***GPAQ (Global Physical Activity Questionnaire) and IPAQ (International Physical Activity Questionnaire) are both validated tools for assessing physical activity which have shown to yield broadly comparable population-level estimates [3].

### References

1. Department of Health (Northern Ireland): Health Survey Northern Ireland: First Results 2023/24. In. Belfast: Department of Health; 2024.
2. Wanner M, Hartmann C, Pestoni G, Martin BW, Siegrist M, Martin-Diener E. Validation of the Global Physical Activity Questionnaire for self-administration in a European context. BMJ Open Sport & Exercise Medicine. 2017;3:e000206. https://doi.org/10.1136/bmjsem-2016-000206
3. Bull FC, Maslin TS, Armstrong T. Global physical activity questionnaire (GPAQ): nine country reliability and validity study. J Phys Act Health. 2009 Nov;6(6):790-804. doi: 10.1123/jpah.6.6.790. PMID: 20101923.

### Supplement 3: Decomposition analysis of inequalities in studied outcomes

#### Table S5: Decomposition of the concentration of four outcomes in Wave 1 (2017/2018)

| Year 2010 (crude data) | Concentration index | % meet PA target | | Time spent in PA | | WEMWBS | | EQ5D-3L | |
|---|---|---|---|---|---|---|---|---|---|
| | | Elasticity | Contribution | Elasticity | Contribution | Elasticity | Contribution | Elasticity | Contribution |
| **Gender** | 0.066 | 0.122 | 0.008 | 0.170 | 0.011 | -0.002 | 0.000 | 0.014 | 0.001 |
| **Age*** | N/A | N/A | 0.048 | N/A | 0.049 | N/A | -0.004 | N/A | -0.001 |
| **Married** | -0.162 | 0.029 | -0.005 | -0.084 | 0.014 | 0.000 | 0.000 | 0.016 | -0.003 |
| **Single** | 0.249 | -0.039 | -0.010 | -0.079 | -0.020 | 0.014 | 0.003 | 0.013 | 0.003 |
| **Education** | 0.189 | 0.080 | 0.015 | -0.076 | -0.014 | 0.022 | 0.004 | 0.017 | 0.003 |
| **Employment** | **0.262** | **0.032** | **0.008** | **0.139** | **0.037** | **0.014** | **0.004** | **0.057** | **0.015** |
| **Long term illness** | -0.267 | -0.341 | 0.091 | -0.103 | 0.027 | -0.025 | 0.007 | -0.119 | 0.032 |
| **Owned housing** | -0.024 | 0.024 | -0.001 | -0.052 | 0.001 | 0.003 | 0.000 | 0.030 | -0.001 |
| **Mortgaged housing** | 0.360 | -0.031 | -0.011 | -0.158 | -0.057 | 0.002 | 0.001 | 0.015 | 0.005 |
| **No car** | -0.445 | 0.035 | -0.015 | 0.031 | -0.014 | -0.014 | 0.006 | -0.008 | 0.004 |
| **Only 1 car** | 0.016 | -0.034 | -0.001 | 0.053 | 0.001 | -0.010 | 0.000 | -0.002 | 0.000 |
| **Deprivation*** | N/A | N/A | 0.016 | N/A | 0.001 | N/A | 0.001 | N/A | 0.003 |
| **Residual** | | | ***-0.007*** | | ***-0.038*** | | ***0.001*** | | ***0.005*** |
| **Total **** | | | **0.137** | | **-0.001** | | **0.022** | | **0.066** |

* Age and deprivation were included as dummy variables; hence, the contribution of these factors is the sum of all sub-groups and single elasticity was only available at sub-group level. ** indicates the overall concentration index of the respective outcomes

**Table S6: Decomposition of the concentration of four outcomes in Wave 2 (2017/2018)**

| Year 2017 (crude data) | Concentration index | % meet PA target | | Time spent in PA | | WEMWBS | | EQ5D-3L | |
|---|---|---|---|---|---|---|---|---|---|
| | | Elasticity | Contribution | Elasticity | Contribution | Elasticity | Contribution | Elasticity | Contribution |
| **Gender** | 0.039 | 0.103 | 0.004 | 0.190 | 0.007 | 0.005 | 0.000 | -0.004 | 0.000 |
| **Age*** | N/A | N/A | 0.042 | N/A | 0.014 | N/A | -0.002 | N/A | -0.001 |
| **Married** | -0.145 | 0.004 | -0.001 | -0.041 | 0.006 | 0.014 | -0.002 | 0.012 | -0.002 |
| **Single** | 0.264 | 0.016 | 0.004 | -0.049 | -0.013 | 0.029 | 0.008 | 0.015 | 0.004 |
| **Education** | 0.188 | 0.046 | 0.009 | -0.227 | -0.043 | 0.013 | 0.002 | 0.026 | 0.005 |
| **Employment** | **0.235** | **0.384** | **0.090** | **0.435** | **0.102** | **0.024** | **0.006** | **0.046** | **0.011** |
| **Long term illness** | **-0.314** | **-0.293** | **0.092** | **-0.126** | **0.040** | **-0.042** | **0.013** | **-0.134** | **0.042** |
| **Owned housing** | 0.027 | 0.046 | 0.001 | -0.003 | 0.000 | 0.012 | 0.000 | 0.021 | 0.001 |
| **Mortgaged housing** | 0.400 | -0.023 | -0.009 | 0.008 | 0.003 | 0.000 | 0.000 | 0.005 | 0.002 |
| **No car** | -0.434 | -0.082 | 0.036 | -0.169 | 0.073 | -0.008 | 0.004 | -0.011 | 0.005 |
| **Only 1 car** | 0.031 | -0.013 | 0.000 | -0.134 | -0.004 | -0.005 | 0.000 | -0.010 | 0.000 |
| **Deprivation*** | N/A | N/A | -0.003 | N/A | -0.047 | N/A | 0.003 | N/A | 0.001 |
| **Residual** | | | ***0.039*** | | ***-0.017*** | | ***0.000*** | | ***0.000*** |
| **Total** | | | **0.303** | | **0.122** | | **0.032** | | **0.066** |

* Age and deprivation were included as dummy variables; hence, the contribution of these factors is the sum of all sub-groups and single elasticity was only available at sub-group level. ** indicates the overall concentration index of the respective outcomes

### Table S7: Decomposition of the concentration of four outcomes in Wave 3 (2023/2024)

| Year 2023 (crude data) | Concentration index | % meet PA target | | Time spent in PA | | WEMWBS | | EQ5D-3L | |
|---|---|---|---|---|---|---|---|---|---|
| | | Elasticity | Contribution | Elasticity | Contribution | Elasticity | Contribution | Elasticity | Contribution |
| **Gender** | 0.083 | 0.142 | 0.012 | 0.114 | 0.009 | 0.006 | 0.001 | 0.014 | 0.001 |
| **Age*** | N/A | N/A | 0.042 | N/A | 0.019 | N/A | -0.005 | N/A | -0.002 |
| **Married** | 0.287 | -0.028 | -0.008 | -0.040 | -0.012 | 0.004 | 0.001 | 0.001 | 0.000 |
| **Single** | 0.217 | -0.072 | -0.016 | -0.072 | -0.016 | 0.004 | 0.001 | 0.010 | 0.002 |
| **Education** | 0.192 | 0.087 | 0.017 | 0.029 | 0.006 | 0.031 | 0.006 | 0.048 | 0.009 |
| **Employment** | 0.205 | 0.287 | 0.059 | 0.275 | 0.057 | 0.035 | 0.007 | 0.106 | 0.022 |
| **Long term illness** | -0.342 | -0.319 | 0.109 | -0.120 | 0.041 | -0.025 | 0.009 | -0.121 | 0.041 |
| **Owned housing** | -0.041 | 0.130 | -0.005 | -0.097 | 0.004 | 0.005 | 0.000 | 0.040 | -0.002 |
| **Mortgaged housing** | 0.350 | 0.098 | 0.034 | -0.070 | -0.025 | 0.007 | 0.002 | 0.013 | 0.005 |
| **No car** | -0.472 | 0.003 | -0.002 | -0.042 | 0.020 | -0.012 | 0.006 | 0.002 | -0.001 |
| **Only 1 car** | -0.030 | -0.087 | 0.003 | -0.034 | 0.001 | -0.005 | 0.000 | 0.000 | 0.000 |
| **Deprivation*** | N/A | N/A | 0.030 | N/A | 0.023 | N/A | 0.002 | N/A | 0.008 |
| **Residual** | | | ***0.060*** | | ***0.002*** | | ***0.005*** | | ***0.005*** |
| **Total** | | | **0.335** | | **0.130** | | **0.034** | | **0.088** |

* Age and deprivation were included as dummy variables; hence, the contribution of these factors is the sum of all sub-groups and single elasticity was only available at sub-group level. ** indicates the overall concentration index of the respective outcomes
